# DEVELOPMENT AND TESTING OF A LOW-COST INACTIVATION BUFFER THAT ALLOWS DIRECT SARS-COV-2 DETECTION IN SALIVA

**DOI:** 10.1101/2021.11.26.21266918

**Authors:** Brandon Bustos-Garcia, Sylvia Garza-Manero, Nallely Cano-Dominguez, Dulce Maria Lopez Sanchez, Gonzalo Salgado Montes de Oca, Alfonso Salgado-Aguayo, Felix Recillas-Targa, Santiago Avila-Rios, Victor Julian Valdes

**Affiliations:** Department of Cell Biology and Development, Institute of Cellular Physiology (IFC), National Autonomous University of Mexico (UNAM), Mexico City, Mexico; Department of Molecular Genetics, Institute of Cellular Physiology (IFC), National Autonomous University of Mexico (UNAM), Mexico City, Mexico; Centre for Research in Infectious Diseases of the National Institute of Respiratory Diseases (CIENI/INER), Mexico City, Mexico; Laboratory of Research in Rheumatic Diseases. National Institute of Respiratory Diseases (INER), Mexico City, Mexico

**Author notes:** Contributed equally (Brandon Bustos-Garcia, Sylvia Garza-Manero).

**Keywords:** SARS-CoV-2, COVID-19 testing, Direct RT-qPCR

## Abstract

Massive testing is a cornerstone in efforts to effectively track infections and stop COVID-19 transmission, including places where good vaccination coverage has been achieved. However, SARS-CoV-2 testing by RT-qPCR requires specialized personnel, protection equipment, commercial kits, and dedicated facilities, which represent significant challenges for massive testing implementation in resource-limited settings. It is therefore important to develop testing protocols that facilitate implementation and are inexpensive, fast, and sufficiently sensitive. In this work, we optimized the composition of a buffer (PKTP) containing a protease, a detergent, and an RNase inhibitor, that is compatible with the RT-qPCR chemistry, allowing for direct testing of SARS-CoV-2 from saliva in an RNA extraction-independent manner. This buffer is compatible with heat-inactivation reducing the biohazard risk of handling the samples. We assessed the PKTP buffer performance in comparison to the RNA-extraction-based protocol of the US Centers for Disease Control and Prevention in saliva samples from 70 COVID-19 patients finding a good sensitivity (82.2% for the N1 and 84.4% for the N2 target, respectively) and correlations (R=0.77, p<0.001 for N1, and R=0.78, p<0.001 for N2). We also propose an auto-collection protocol for saliva samples and a multiplex reaction to reduce the number of PCR reactions per patient and further reduce overall costs while maintaining diagnostic standards in favor of massive testing.

## INTRODUCTION

Despite vaccination strategies against the Severe Acute Respiratory Syndrome Coronavirus 2 (SARS-CoV-2) successfully advancing in many countries, the appearance of variant lineages with higher viral-titers or greater transmission rates, such as B.1.617 (delta variant), has severely impacted the progress made thus far to control SARS-CoV-2-associated disease (COVID-19) even on vaccinated individuals [1-4]. As COVID-19 confirmed cases continue to increase in countries facing recent waves of infection, massive testing campaigns constitute a key strategy to trace and stop the virus spreading, while vaccination continues. On the other hand, a possible outcome of the SARS-CoV-2 pandemic is that it will eventually become a seasonal health concern [5], which further highlights the relevance of developing safer, cheaper, faster and more feasible universal testing strategies that may act as a surveillance approach in working environments and schools [6].

To this day, the gold-standard for SARS-CoV-2 diagnosis is the detection of the viral genetic material by real-time RT-PCR, which involves: 1) collection of nasopharyngeal swabs (NPS) by specialized personnel, 2) purification of the viral genetic material with commercial kits, and 3) detection of at least two viral targets and a human target, usually RNAse P (hRP), by RT-qPCR into three separate reactions. These requirements bear several drawbacks if massive testing is expected, such as: i) personal protective equipment (PPE) and specialized personnel are required to collect and process the sample, ii) commercial RNA extraction kits are expensive and their supply has been limited during the pandemic, and iii) detecting targets into separate reactions results in the need of higher amounts of reagents (and higher costs). All of these constitute limiting factors for massive testing.

The use of simpler-to-collect specimens for detecting SARS-CoV-2 supported by alternative processing procedures could aid to overcome some of the limitations associated to NPS [7, 8]. Raw saliva has been proven a reliable source of viral genetic material as viral loads in this fluid can be equivalent to NPS [8-11]. Several processing-sample alternatives have been reported for the use of saliva as specimen for SARS-CoV-2 infection diagnosis, including direct lysis by heating to avoid the RNA extraction step [12, 13] or using commercial reagents [10], common buffers such as PBS or TE [13], Proteinase K [14, 15] or guanidine hydrochloride [16] as lysing-inactivating solutions. However, some of these strategies have been reported to inhibit the RT-qPCR reaction to some degree [13, 17], showing varying sensitivity for detecting viral targets in saliva. Thus, in most countries, RNA extraction-free protocols in saliva have not been considered to fully substitute COVID-19 diagnostics and further systematic studies are needed.

In this work, we systematically tested several buffer formulations and optimized Proteinase K and detergent concentrations to identify the conditions that, in conjunction to the addition of a broad-range RNAse inhibitor, improved viral genetic material detection in saliva by direct RT-qPCR without nucleic acid extraction. This optimized buffer (PKTP) allows an easy sample inactivation protocol and circumvents the RNA extraction step, dramatically reducing processing time, costs and biohazard risk. Additionally, PKTP can be used in several workflows, including standard RNA purification using commercial kits. We validated the use of PKTP buffer on 70 COVID-19 patients by comparing its performance vs. the CDC-validated RNA-extraction protocol on saliva samples. To facilitate the use of PKTP buffer, we propose a self-collecting sample strategy based on the use of graduated microcapillary tubes to pour saliva into PCR tubes containing PKTP buffer, which allows direct heat-inactivation of the sample completely removing the need of manipulation of infectious specimens by laboratory personnel. To further reduce testing costs, we optimized a multiplex probe set to simultaneously detect two different viral genes and a human endogenous target in a single reaction.

## RESULTS

### Detergent screening for direct SARS-CoV-2 detection in saliva by RT-qPCR

Viral-titers in saliva have been reported to be equivalent or even higher than those in nasopharyngeal swabs from COVID-19 patients [9-11]. Therefore, we aimed to design a buffer formulation that allowed direct detection of SARS-CoV-2 by RT-qPCR in saliva samples avoiding RNA isolation, biohazard risks and reducing processing times. Solutions containing proteases only have been used to lyse virus in saliva samples [15], thus our starting point was a Proteinase K solution [800 µg/mL] in calcium- and magnesium-free phosphate saline buffer (PK) supplemented with different detergents at 0.25 % to improve buffer formulation. We mixed SARS-CoV-2 positive saliva with PK buffer and different detergents prior to heat-inactivation of the sample. Then, 4 µL were directly placed in two individual RT-qPCR reactions to detect the SARS-CoV-2 E gene and the human RNase P gene (hRP), as endogenous control. hRP detection was efficient in all formulations except for cetrimonium bromide (CTAB). N-Laurylsarcosine and Na-Deoxycholate showed the highest efficiency with a 100-fold increase vs. PK alone, followed by Tween 80 with a 10-fold change (Fig. 1A, left). On the other hand, most detergents inhibited detection of the SARS-CoV-2 E gene except Tween 80, N-Laurylsarcosine and SDS. Noteworthy, Tween 80 improved SARS-CoV-2 E gene detection by 10-fold in comparison with PK alone (Fig. 1A, right). Since PK + Tween 80 (henceforth referred to as PKT) was the sole combination that improved detection efficiency for both targets, we chose this combination for further optimization.

**Figure 1.**
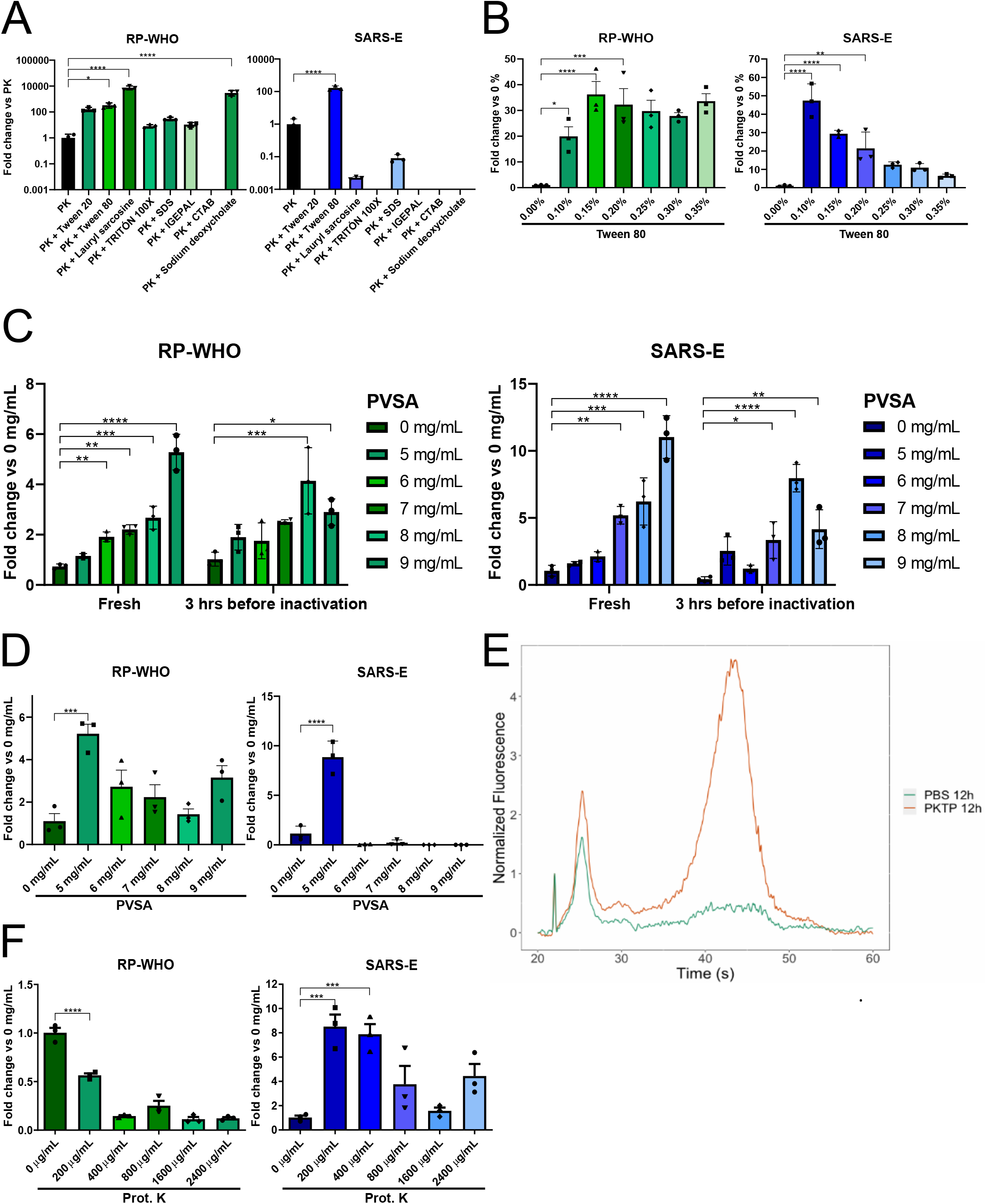
PKTP buffer allows direct SARS-CoV-2 detection by RT-qPCR protecting saliva from degradation. COVID-19 positive saliva was mixed in a 2:1 ratio with buffer and heat-inactivated (10 min at 96 ºC). Direct RT-qPCR detection of human RP (green) and viral E (blue) targets using CDC-designed probes was performed in 10 µL reactions. **A)** Different detergents at 0.25 % were screened for human and viral RNA detection. Signal was normalized to DPBS + Proteinase K (PK) only. **B)** Detection efficiency for both targets using PK buffer with different Tween 80 concentrations (PKT) normalized to PK only. **C)** Incremental effect of PVSA concentration on detection of both targets in freshly inactivated samples versus samples incubated 3 hours at room temperature with PKTP buffer before heat inactivation. **D)** Effect of PVSA concentration after heat inactivation. **E)** RNA size distribution frequency in saliva samples incubated at 4 ºC with PKTP or PBS alone assessed by Bioanalyzer. **F)** Proteinase K titration curve with full PKTP buffer formulation to favor viral target detection. Data were compared using a one-way ANOVA followed by Dunnett’s multiple comparisons test; (*, p = 0.05), (**, p = 0.01), (***, p = 0.001), (****, p = 0.0001). Bars represent means and 95 % confidence intervals.

To determine the precise detergent amount, we performed a concentration curve of Tween 80 in PK buffer. We mixed SARS-CoV-2 positive saliva samples with PK buffer containing different concentrations of Tween 80, heat-inactivated the samples and assessed each target individually. In the case of hRP, Tween 80 concentrations ranging from 0.15 % to 0.35 % resulted in a 30-to-40-fold increase in detection (Fig. 1B, left), while SARS-CoV-2 E gene detection improved as Tween 80 concentration diminished, with a peak of a 40-fold increase at 0.1 % (Fig. 1B, right). Although the most efficient condition for hRP was 0.15 % Tween 80, we selected 0.1 % to favor the detection of viral genetic material since differences between 0.15 % and 0.1 % are not statistically significant for hRP.

### Addition of an RNase inhibitor enhances viral genetic material detection in saliva

Saliva is a source of RNases that may degrade the SARS-CoV-2 RNA genome, compromising its detection by RT-qPCR [18]. To overcome this situation, we added the thermostable RNase inhibitor Poly-Vinyl Sulfonic Acid (PVSA) to the PKT buffer (hereafter PKTP), a low cost-reagent that can enhance RNA preservation at low concentrations [19]. We performed a PKT+PVSA concentration curve and compared the detection efficiency between freshly inactivated samples and samples that were mixed with PKTP buffer and incubated 3 hours at room temperature before heat inactivation to test for RNA stability during bench-work time prior to sample processing. hRP detection was significantly increased in a PVSA dose-dependent manner, both with fresh and 3-hour samples (Fig. 1C, left). Similarly, SARS-CoV-2 E detection displayed a 10-fold increase in fresh lysates at 9 mg/mL of PVSA (Fig. 1C, right), suggesting that higher concentrations of PVSA are advantageous. Next, to evaluate the stability of the RNA after inactivation we repeated the PVSA concentration curve incubating the samples for 3 hours after heat inactivation. This time, hRP detection was more efficient as PVSA concentration decreased, with a peak at 5 mg/mL (Fig. 1D, left), while SARS-CoV-2 E was almost undetectable at higher concentrations of PVSA (Fig. 1D, right). It has been suggested that PVSA may inhibit RT-qPCR at high concentrations presumably as a consequence of its nucleic acid mimicking characteristics [15, 19]. However, the use of 5 mg/mL of PVSA displayed higher efficiency than PKT alone (Fig. 1D, right). We concluded that this concentration of the RNase inhibitor is optimal for the detection of both the endogenous and the viral targets by RT-qPCR on saliva protecting the viral RNA before and after heat inactivation of the sample. To further assess the stability of RNA, saliva was mixed with PBS or PKTP (2:1) and incubated overnight at 4 ºC, then total RNA integrity was assessed using a Bioanalyzer instrument. The resulting electropherogram showed that the RNA in PKTP buffer was enriched in high molecular weight RNA molecules that are absent when saliva is incubated in PBS alone (Fig. 1E), indicating that PKTP buffer is able to protect the RNA present in saliva for at least 12 h at 4 ºC.

Finally, as proteinase enzymatic activity may be perturbed by the new buffer composition, we performed a last titration curve of Proteinase K. Detection of hRP was significantly increased at lower concentrations of Proteinase K (Fig. 1F, left). In contrast, that of SARS-CoV-2 E was clearly improved by Proteinase K addition at all concentrations, with the highest efficiency at 200 µg/mL (Fig. 1F, right). Since this WHO-designed hRP primer set does not discriminate between gDNA and mRNA, we speculated that Proteinase K may particularly affect gDNA detection, while promoting a more efficient viral RNA release from viral particles contained in the saliva samples. As differences in hRP detection between 0 µg/mL and 200 µg/mL accounted for only a 0.5-fold decrease (about one single Ct unit), we set 200 µg/mL as the final concentration of proteinase K, to favor viral target detection.

### Optimization of saliva lysis conditions

Once we defined the optimal buffer formulation for the direct detection of a viral gene in saliva by direct RT-qPCR, we assessed mixing saliva with PKTP buffer at different ratios: 1:1, 2:1 and 3:1. Although the 1:1 ratio worked better for hRP detection, this condition displayed the greatest variation among replicates (Fig. 2A, left). Otherwise, there were no significant differences in SARS-CoV-2 E gene detection among all tested ratios (Fig. 2A, right). Hence, we chose the 2:1 saliva:buffer ratio as it was the most robust condition to lyse saliva samples in PKTP buffer.

**Figure 2.**
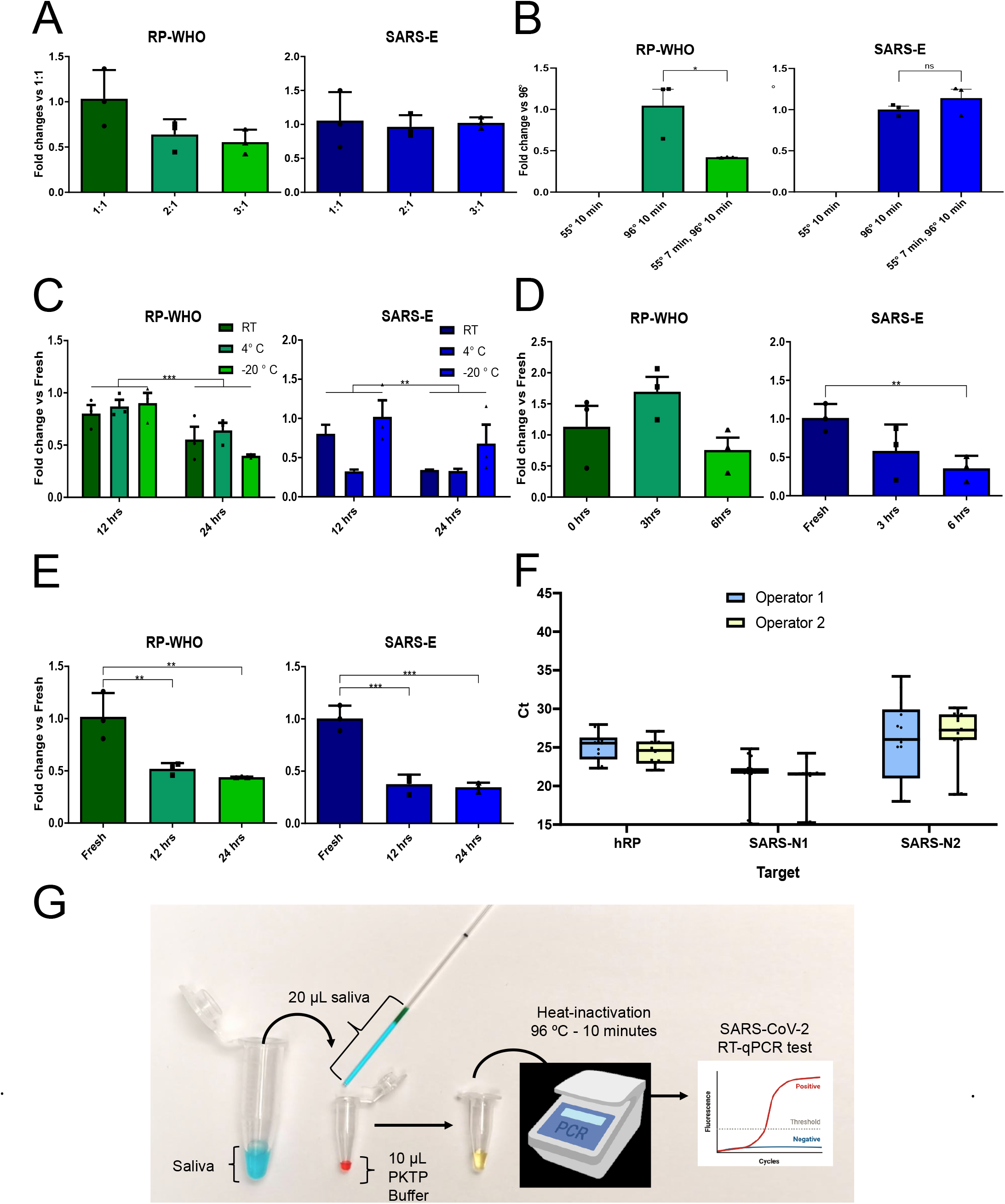
PKTP-based protocol optimization. Detection of hRP (green) and viral SARS-E (blue) targets by RT-qPCR assessed on COVID-19 positive saliva. **A)** Optimization of saliva:buffer mix ratio with PKTP buffer as lysing solution. **B)** Optimization of the heat-inactivating program. **C)** PKTP buffer stability determination by using buffer stored at different temperatures for twelve and twenty-four hours. **D)** Stability tests of samples three and six hours before PKTP heat-inactivation stored at room-temperature. **E)** Stability test of samples frozen for twelve and twenty-four hours after heat inactivation stored at – 20 ºC. **F)** Repeatability and reproducibility tests of PKTP-protocol. **G)** Proposed self-applicable saliva obtention strategy using graduated microcapillary tubes to minimize biohazard risk. Data were compared using a one-way ANOVA followed by Dunnett’s multiple comparisons test except for F); (*, p = 0.05), (**, p = 0.01), (***, p = 0.001), (****, p = 0.0001). Bars represent means and 95 % confidence intervals. Biorender was used to create a part of the illustration.

Next, as Proteinase K displays its peak-activity at 55-65º C, we aimed to identify the best inactivating conditions by testing three different inactivating programs: 1) 10 min at 55° C, 2) 10 min at 96° C, and 3) 7 min at 55° C followed by 10 min at 96° C. hRP was clearly detectable with programs 2 and 3, although program 2 showed a significantly higher detection efficiency (Fig. 2B, left). In contrast, SARS-CoV-2 E detection worked similarly between programs 2 and 3, with a slight improvement in program 3 (Fig. 2B, right). The first program resulted in no detection of any target (Fig. 2B), probably because non-inactivated Proteinase degrades the Taq polymerase and reverse transcriptase enzymes. Thus, we established 10 min at 96° C as the simplest working conditions for inactivating saliva samples in PKTP buffer.

Additionally, we assessed the stability of the PKTP buffer by storing it at different temperatures. We stored aliquots from the same batch at room temperature (RT), 4° C and -20° C during twelve and twenty-four hours. The stored buffer aliquots were used to inactivate SARS-CoV-2 positive saliva samples and the RT-qPCR reactions were run in parallel vs. a fresh buffer batch. hRP detection resulted in a slight 0.2-fold decrease at twelve hours in all temperatures, and approximately a 0.4-fold decrease (about one single Ct unit) at twenty-four hours, when compared to the fresh buffer batch (Fig. 2C, left). However, storage temperature differences did not reach statistical significance (p = 0.68 and 0.36, respectively). In contrast, SARS-CoV-2 E detection showed a marked 0.7-fold decrease (although not significant, p = 0.086) when the buffer was stored at 4° C, even for twelve hours, while storing the buffer at -20° C resulted in no significant decrease (p = 0.478) at twelve hours and a 0.3-fold decrease at twenty-four hours (Fig. 2C, right). Notably, both hRP and SARS-E showed a statistically significant decrease at twenty-four hours when compared to twelve-hour-incubated samples at all temperatures (p= 0.0004 and 0.0347, respectively). Altogether, these observations suggest that human and viral genetic materials display different stability profiles. We concluded that the most convenient temperature to store the PKTP buffer is -20° C, as viral detection is more robust in this condition.

Next, we evaluated the stability of the saliva:buffer mix before and after heat-inactivation to define the versatility of the use of PKTP buffer in different workflows. First, we performed a stability assay of saliva:buffer mix for three and six hours at room temperature before heat-inactivation. hRP detection showed no statistical differences among the different conditions, although there was an apparent 0.3-fold decrease (not significant, p = 0.24) at six hours (Fig. 2D, left). In contrast, SARS-CoV-2 E detection displayed a trend to decrease over time, with a significant 0.6-fold decrease (p = 0.0097) at six hours (Fig. 2D, right). Additionally, we checked the stability of saliva:buffer mix after heat inactivation for twelve and twenty-four hours stored at -20º C. In these experiments, both human and viral targets showed a significant 0.5-fold decrease at twelve and twenty-four hours (Fig. 2E, p = 0.001), which seems to be related to freezing-thawing cycles of the sample rather than the buffer [13]. In conclusion, the PKTP buffer can be introduced in different workflows, including storage before and after heat inactivation of samples, although a single Ct unit decrease is to be expected when compared to freshly inactivated samples.

Finally, we tested sample processing with PKTP for both repeatability and reproducibility by testing intra-run replicates and intra-operator variability. We prepared aliquots from each of five SARS-CoV-2 positive saliva samples to ensure both operators worked with the same samples and identical numbers of freezing-thawing cycles. Operators in two separate facilities performed three technical replicates (i.e. repeatability) for each PKTP-inactivated sample using the standard CDC-detection probes N1, N2 and hRP. Later, on a different day, each operator thawed samples right before the experiment and processed them using the PKTP buffer as described above (i.e. reproducibility). In all cases technical replicates yielded precise results (Fig. 2F). Furthermore, there was no statistical difference in results for any of the three targets between operators (Fig. 2F, p = 0.464), as determined by a two-way ANOVA test. Altogether, our results indicate that the PKTP buffer can be used by different technicians obtaining identical results, and that its use can be introduced into different working routines.

### Saliva self-collection protocol

To further reduce the biohazard risk and to consider low-budget settings where no PPE nor trained personnel are available to collect samples, we developed a saliva self-collection strategy using the PKTP buffer. A commercially graduated microcapillary tube is used to place 20 µL of saliva into the tube containing PKTP buffer as represented in Fig. 2G. The microcapillary tube has a mark at 20 µL (green line) and patients are instructed to directly pour saliva up to this mark. Then, the saliva contained in the microcapillary tube is placed into a regular PCR tube by the patient, containing 10 µl of PKTP buffer to guarantee the 2:1 saliva:buffer ratio. This mix is then incubated for 10 min at 96º C in a thermal cycler for heat-inactivation of the samples, allowing subsequent RT-qPCR without the need of pipetting, significantly reducing biohazard risk during sample processing.

### Use of PKTP buffer in a clinical setting

We evaluated the performance of our optimized PKTP inactivation protocol which allows direct qPCR detection of SARS-CoV-2 in saliva without the need to purify RNA, in a hospital setting. This was assessed on 70 frozen saliva samples from previously diagnosed COVID-19 patients at the National Institute of Respiratory Diseases (INER), a tertiary referral hospital in Mexico City. We divided each stored sample into two: one half was heat-inactivated in PKTP buffer and the other half was processed by the standard RNA-extraction-based protocol from the US Centers for Disease Control and Prevention (CDC). RT-qPCR reactions were run in parallel to detect viral N1 and N2, and human hRP targets into separate reactions using the CDC-designed probes to compare sensitivity between the two protocols. hRP was detected in all samples, showing a median increase of 3.5 Ct units when using the PKTP protocol (Fig. 3A, top panel). Differences in the amount of template added to the reactions between protocols (6 times higher when RNA extraction is performed) may have accounted for the delay observed in the amplification plot. Similarly, Cts from viral genes from the purified RNA were lower than those from lysed saliva samples, possibly due to the aforementioned difference in template amount, with a mean difference of 3.64 Cts (N1) and 8.32 Cts (N2). To correct for the difference in template amount between protocols, the Ct values form PKTP were adjusted by the dilution factor showing a Ct difference of 0.91, 1 and 5.7 for hRP, N1 and N2, respectively. Overall, both N1 and N2 viral targets were detected in approximately 80% of the PKTP-processed samples (Fig. 3B and C, top panels): 60 and 61 samples were detected as SARS-CoV-2 positive with the N1 and N2 probes. This represents a diagnostic sensitivity of 82.22% (67.95%-92%) for the N1 probe and 84.44% (70.54%-93.51%) for the N2 probe using non-adjusted PKTP values.

**Figure 3.**
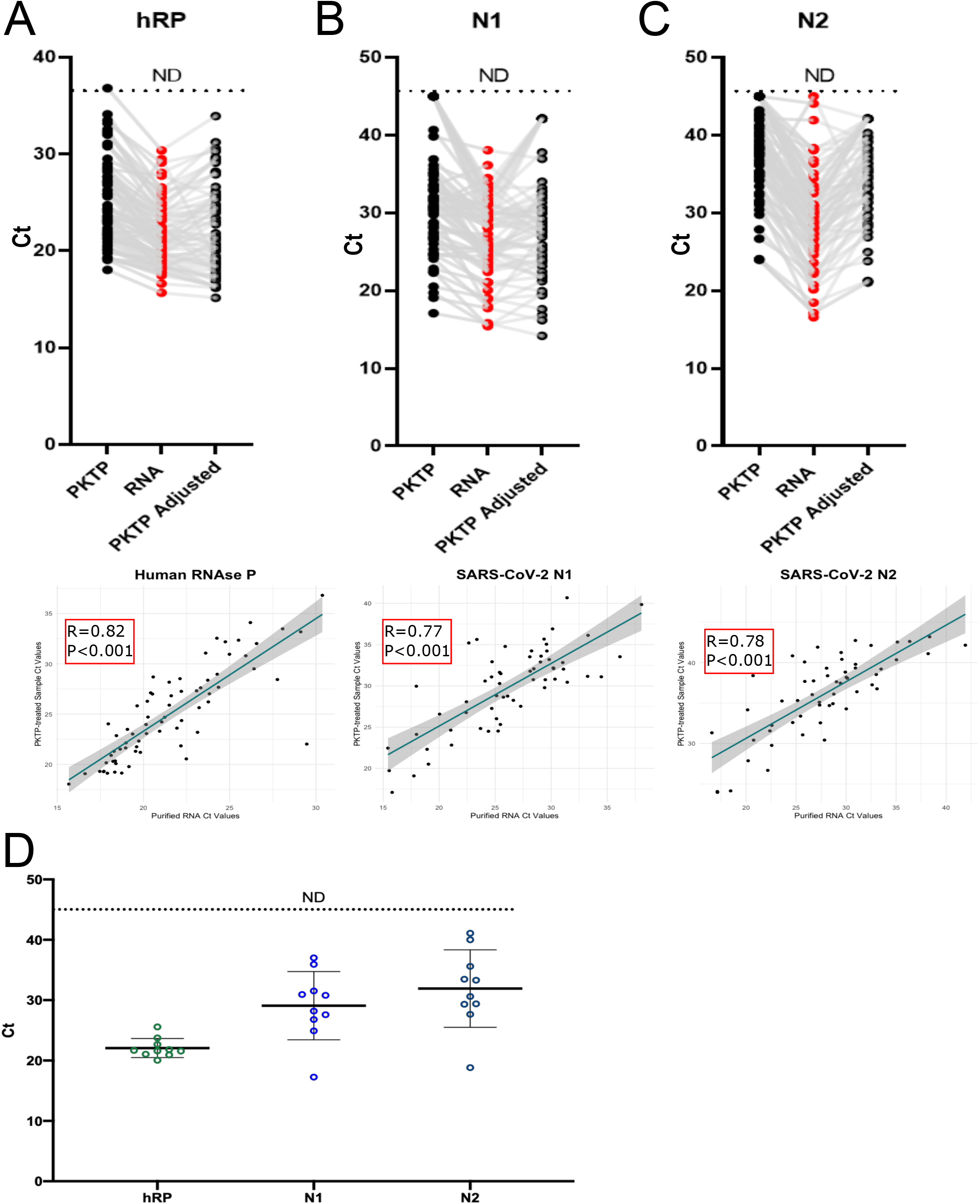
PKTP protocol performance in a clinical setting. Frozen saliva from 70 previously diagnosed COVID-19 patients at the INER hospital in Mexico City were used to compare human and viral RNA detection between the CDC RNA-extraction protocol (RNA) and our PKTP-inactivating protocol followed by direct RT-qPCR without RNA extraction. Reaction volume was 20 µL. PKTP adjusted: the 6-fold template difference was mathematically corrected. **A)** hRP FAM, **B)** SARS-N1 FAM and **C)** SARS-N2 FAM Cts comparing protocols using paired samples (top panels). Lower panels display Pearson’s correlation test between protocols for each target. n = 70. **D)** Ct values obtained by purifying RNA with a commercial kit (Qiagen) from PKTP-inactivated saliva samples.

We performed Pearson correlation analyses comparing the Cts of purified RNA vs. PKTP non-adjusted values. A good correlation was found with the three CDC-probes (Fig 3A, 3B and 3C, bottom panels); Cts obtained using the hRP probe had a correlation coefficient of 0.82, while the correlation coefficients of Cts from N1 and N2 were 0.77 and 0.78, respectively. Our results show that saliva samples lysed with the PKTP buffer have a similar detection rate, although with a slightly lower performance in the RT-qPCR reactions using CDC-designed probes, compared to the standard RNA purification-based protocol from saliva samples.

An increase in Cts when using the PKTP protocol could affect diagnostic sensitivity especially when samples display low viral titers (> 30 Cts), thus we tested if our buffer could also be compatible with downstream RNA extraction using commercial kits. To address this, ten COVID-19-positive saliva samples with varying Ct values were heat-inactivated in PKTP buffer, their RNA was extracted with a CDC approved kit [20], and the viral targets were assessed using the same CDC-designed probes. All target genes were detected except for N2 in one low-titer sample (Fig. 3D). This result suggests that the PKTP buffer can be used to inactivate saliva samples for further RNA extraction with commercial kits, pointing out the possibility to re-process suspicious samples when viral titers are too low to be detected by the direct PKTP-to-qPCR protocol.

### Reducing the number of RT-qPCR reactions by multiplexing

The standard CDC and WHO protocols use two different sets of probes for detecting the SARS-CoV-2 genetic material. The CDC N1 and N2 probes target the nucleocapsid gene N, while the WHO probes test for two independent genes: the envelope gene E and the RNA dependent RNA polymerase gene RdRP. In addition, both protocols target RNase P (hRP) as an endogenous control, requiring three independent RT-qPCR reactions per sample to emit a result. Hence, we aimed to reduce the number of reactions per sample to further reduce costs and allow more samples to be processed within the same RT-qPCR run by using various probes with different fluorophores within a single multiplex reaction. To achieve this, we first evaluated the performance of the CDC and WHO probes labeled with common fluorophores for the detection of SARS-CoV-2 genetic material. We used as templates serial dilutions of RNA extracted from saliva samples of three COVID-19 patients yielding different viral titers. We observed that the SARS-CoV-2 RdRP gene was consistently amplified at higher Cts than the N and E viral genes, which resulted in a lower sensitivity to detect low viral titers (Fig. 4G). In contrast, the N1 and N2 probes showed the lowest Ct values in each condition (Fig. 4A-C). The E gene probes showed similar kinetics to the N probes in most of the tested dilutions (Fig. 4D-F). Thus, we selected N1 and E as viral targets to integrate the multiplex reaction in favor of testing two different viral genes. Amplification plots of these two viral targets showed consistency along serial dilutions of the sample RNA, while Cts exhibited linearity according to the viral titer (Fig. 4). However, we observed that the sensitivity to detect the E gene at low viral titers was low compared to N1 and N2 (Fig. 4), highlighting the relevance of testing more than one viral target in the diagnosis of SARS-CoV-2.

**Figure 4.**
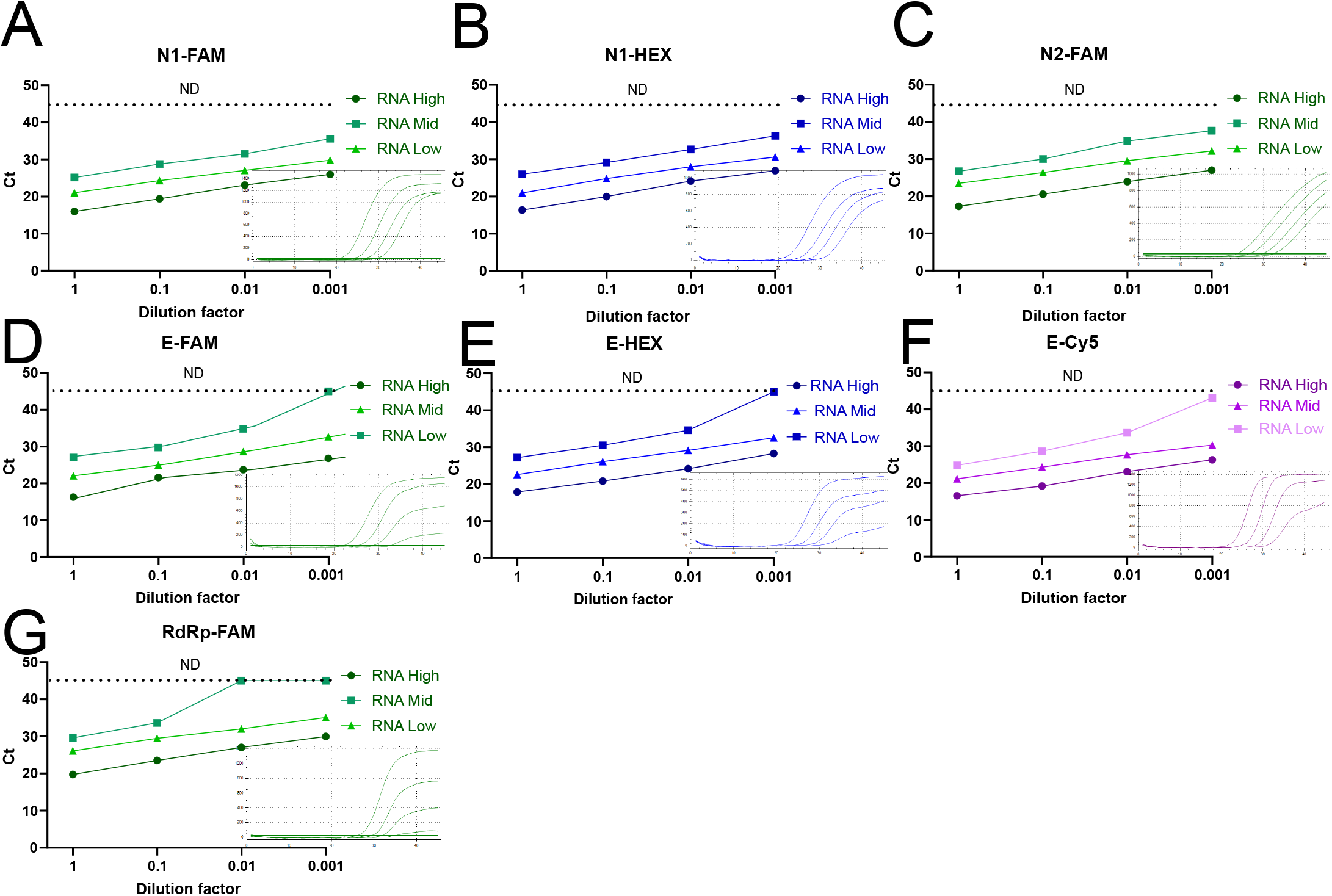
Viral targets detection efficiency. RNA from three different SARS-CoV-2 viral-load saliva samples (low, medium, high) was extracted and tested by RT-qPCR at serial dilutions. Ct values of each singleplex reaction using probes labeled with different fluorophores against viral targets N1, N2, E and RdRP are shown in the graphs. Amplification plot of the serial dilutions corresponding RNA with medium viral load are included for each target.

Noteworthy, the E probe labeled with Cy5 detected low viral titers that were not detected when the probe was marked with other fluorophores (Fig. 4D-F), raising the possibility that different fluorophore combinations could influence performance of the RT-qPCR. To explore this, we designed two different multiplex reactions. In both, the highly sensitive CDC N1 probe labeled with FAM was included (reference probe). For multiplex 1 we used the E-Cy5 and hRP-HEX probes, and for multiplex 2 we used the E-HEX and hRP-Texas Red probes. We contrasted the performance of each probe from the multiplex 1 and 2 reactions with that of the corresponding singleplex reactions, using as templates the same serial dilutions of RNA extracted from saliva samples of three COVID-19 patients with different viral loads. There were no marked differences in the sensitivity, nor in the Cts, when the mentioned probes were used in singleplex or in multiplex reactions (Fig. 5). In agreement with previous results, the E-Cy5 probe displayed higher sensitivity than the E-HEX probe in all conditions, providing similar Cts than the N1-FAM probe (Fig. 4 and 5A).. However, both multiplex combinations generated the expected amplification plots and efficiently detected the three targets (Fig. 5A).

**Figure 5.**
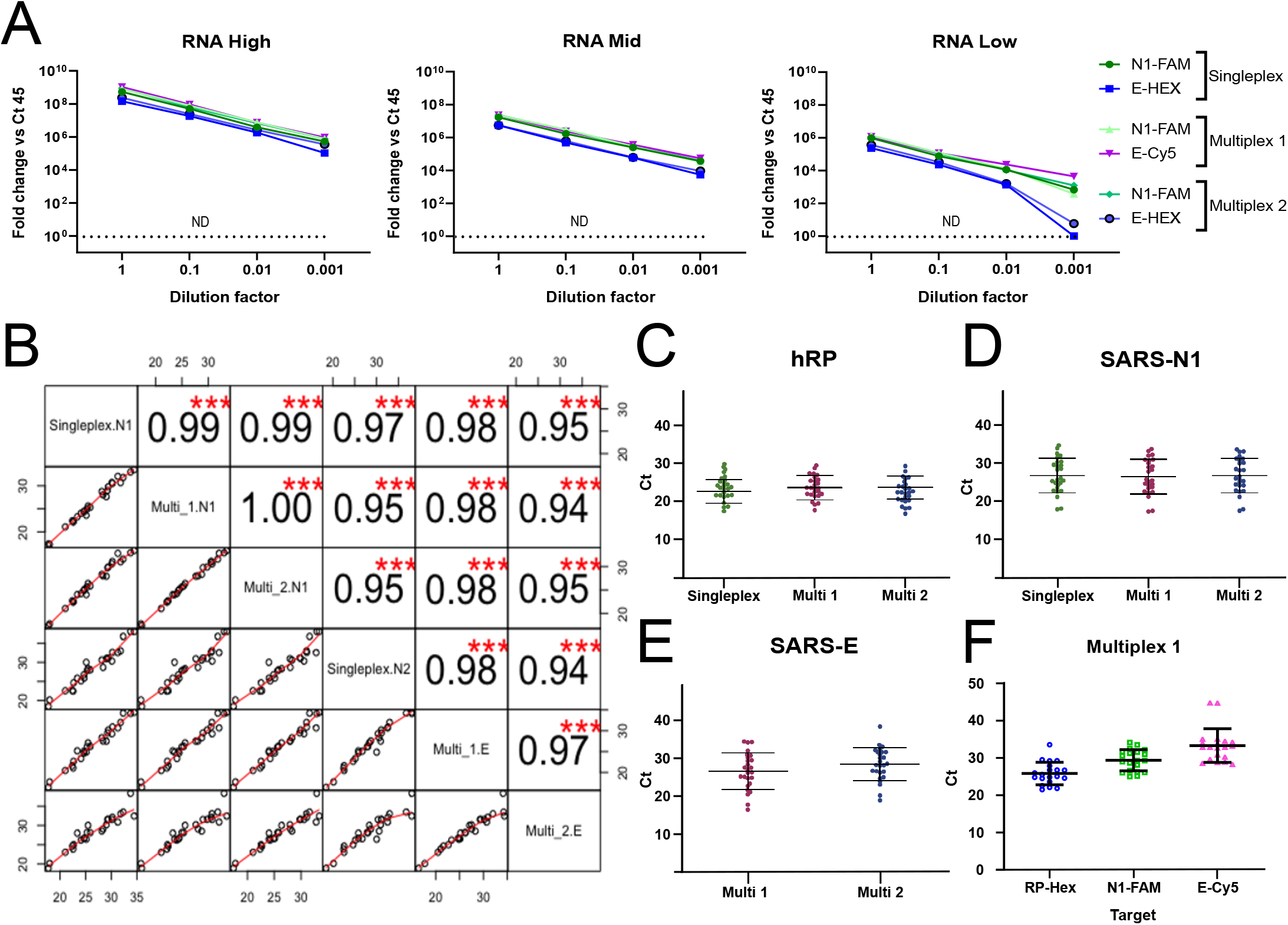
Multiplex reaction setting. **A)** RNA from three different SARS-CoV-2 viral-load saliva samples (low, medium, high) was extracted and tested by RT-qPCR at serial dilutions. Graphs show the viral target detection efficiency in singleplex reactions in comparison to two multiplex reactions for three targets at a time. **B)** Frozen saliva from 26 previously diagnosed COVID-19 patients at the INER hospital in Mexico City were used to compare the RT-qPCR signal between the CDC probes in singleplex and the multiplex reactions in the RNA-extraction protocol. Pearson correlation coefficients of Cts obtained for each target using singleplex or multiplex reactions. **C)** hRP, **D)** SARS-N1 and **E)** SARS-E Cts. **F)** Multiplex-1 Cts obtained for each target from 20 PKTP-inactivated samples without RNA-extraction.

Next, we evaluated the performance of multiplex 1 and 2 in a larger group of saliva samples from previously-diagnosed COVID-19 patients enrolled at the INER Hospital in Mexico City and compared it with that of the original singleplex RNA extraction-based CDC protocol. We used 26 stored samples, with a wide-range of initial viral titers (Cts from 15 to 35 for CDC N1 probe). Cts obtained with multiplex formulation 1 and 2 showed excellent correlations with Cts obtained from singleplex reactions (Fig. 5B). Furthermore, all the viral probes showed high correlation between them, regardless of their use in singleplex or multiplex reactions (Fig. 5B, Pearson correlation coefficients from 0.94 to 0.999). As expected, when reaction efficiencies are identical, there were no significant changes in Cts obtained between the three methods when using the RP probe (Fig. 5C, p > 0.05) nor the N1 probe (Fig. 5B and D, p > 0.05). These results suggest that viral gene detection is equally efficient using singleplex and multiplex tests. On the other hand, the E-Cy5 and E-HEX probes showed an excellent correlation (Fig. 5B), although there is a mean difference of 1.8 Ct (E-Cy5 mean Ct = 26.6 (24.6-28.6), E-HEX mean Ct = 28.4 (26.6-30.2), p = 0.16, Fig. 5E), which could possibly be due to differences in fluorescence detection efficiency of the two fluorophores in the system (different quantum efficiency in the detector, filter efficiency, etc). Therefore, multiplex 1 was chosen for successive experiments.

Finally, we investigated whether the chosen multiplex 1 was compatible with the PKTP buffer by testing 20 frozen COVID-19 positive saliva samples that were previously heat-inactivated in PKTP buffer. The viral targets N1-FAM and E-Cy5 as well as hRP-HEX were evaluated by RT-qPCR in the same reaction (Fig. 5F). All targeted genes were detected in the PKTP-inactivated samples, although the Ct means were higher than those observed when using the multiplex 1 set on extracted RNA (Fig. 5C, D, E, F). These results demonstrate that two viral genes and one endogenous control can be detected in a single RT-qPCR reaction, using saliva samples without the need to isolate RNA, reducing time, reagents, costs and biohazard risks.

## DISCUSSION

Epidemiological surveillance of SARS-CoV-2 will be especially important even in the vaccinated population as worldwide recent evidence has revealed [1, 2, 4, 21]. In this work, we optimized the composition of a buffer (PKTP) that is compatible with the RT-qPCR chemistry, allowing for direct SARS-CoV-2 testing in saliva without RNA extraction. This buffer is compatible with a heat-inactivation step that has been previously demonstrated to eliminate SARS-CoV-2 in saliva [13, 22], minimizing the biohazard risks of handling the samples. However, RNA degradation is a concern when heating or storing samples, as saliva contains thermo-stable RNases that may compromise the diagnosis [18]. Our own data showed that storage of saliva resulted in low molecular weight RNA molecules indicative of degradation (Fig. 1E), thus we included PVSA, a low-cost thermo-stable RNase inhibitor, in our buffer. Addition of PVSA resulted in an RNA size distribution enriched with larger fragments (Fig. 1E) when storing saliva before heat-inactivation and improved amplification of the E viral gene in a dose-dependent manner. However, after heat-inactivation, high concentrations of PVSA seemed to inhibit viral target detection while cellular genes are less affected (Fig. 1D). Interestingly, our proteinase K and detergent concentration screenings showed that cellular and viral genes have different behaviors: most detergents allowed amplification of hRP but only Tween 80 at low concentration improved viral E gene amplification (Fig. 1A-B). An opposite trend was observed with proteinase K as higher concentration decreased hRP detection while the viral target was more resilient (Fig. 1F). These results suggest that viral RNA has particular characteristics that are not equivalent to the endogenous transcripts, which should be taken into consideration when developing novel testing strategies for coronaviruses. Overall, addition of PVSA, Tween 80 and Proteinase K at the optimized concentrations improved RT-qPCR detection of cellular and viral targets genes before and after inactivation of saliva (Fig. 1).

Freezing saliva is a major cause of RNA degradation accounting for an increase of 2-3 Cts when detecting SARS-CoV-2 RNA by qPCR [13]. Of note, the freezing effect on PCR performance of saliva mixed with PKTP was nearly a 0.5 Ct increase (Fig. 2E). Consistently, our stability tests showed that viral and cellular targets lose only ∼0.5 Ct units when stored overnight at -20º C after heat-inactivation, or 3h at RT before inactivation (Fig. 2D and E). Thus, samples can be stored frozen before and after heat-inactivation with less than a Ct unit increase. On the other hand, a stability test of the PKTP buffer itself showed that when frozen, storage does not compromise reaction performance (Fig. 2C). Therefore, PKTP use is suitable for different working routines without important performance differences to be expected. Additionally, a self-applicable saliva obtention strategy using graduated microcapillary tubes (Fig. 2G) could allow patients to directly deliver 20 µL of their own saliva into PCR tubes containing 10 µL PKTP buffer that can be directly heat-inactivated in only 10 minutes with no further manipulation before qPCR. This significantly reduces the biohazard risk and the need of trained personnel, PPE or dedicated facilities for RNA extraction.

Validating the use of the PKTP buffer in a clinical setting, paired comparisons between the CDC RNA extraction-based protocol and our PKTP buffer method in saliva samples from a group of 70 COVID-19 patients showed that hRP was detected in all samples, while N1 and N2 were detected in approximately 80 % of samples when using the PKTP protocol. This represents a diagnostic sensitivity of 82.22% (67.9%-92%) for the N1 probe and 84.44% (70.54%-93.51%) for the N2 probe (Fig. 3B). Importantly, Cts in PKTP-processed samples seemed to be higher than those obtained for isolated RNA (Fig. 5C, D, E, F). Differences in the template amount included in the reactions between protocols may account for the delay observed in the amplification plot: while 3.33 µL of saliva were included per reaction when using the PKTP inactivation protocol, the RNA purification method allows to resuspend the RNA in a smaller volume and include the equivalent of 20 µL of saliva in the RT-qPCR. Indeed, correction of the Ct values by the dilution factor, account for ∼1 and ∼5 Cts average differences for N1 and N2, respectively. Furthermore, as PKTP-inactivated samples are compatible with standard RNA-extraction kits (Fig. 3D), this may compensate for differences in initial template amounts while still minimizing the biohazard risk of purifying RNA from saliva in specific clinical settings. Nevertheless, the decrease in sensitivity and the gain in simplicity and cost-effectiveness should be weighed for implementation purposes.

Finally, to further simplify the protocol, increase the number of samples that can be processed at the same time, diminish the amounts of reagents required per sample and reduce costs, we optimized multiplexed reactions to detect the three targets in one RT-qPCR reaction and showed that multiplexing is compatible with the PKTP buffer (Fig. 5F). For the multiplex reaction, we chose viral genes N1 and E as our data showed that these targets were more sensitive and reliable in comparison to RdRP on purified RNA from samples with different viral loads (Fig. 4). Nevertheless, the method could be adapted to include other probe combinations or even to test for other pathogens that may be relevant in the case of co-infection of SARS-CoV-2 and influenza virus [23, 24].

Overall, our results showed that the protocol based on the PKTP buffer provides a good alternative to minimize the biohazard risk of handling COVID-19 samples, eliminating the need of trained personnel, PPE or dedicated facilities to perform the RNA extraction. In our hands, false positive results were virtually nonexistent when assaying two viral targets, making the PKTP buffer an excellent alternative for massive testing in saliva not only in settings where trained personnel is scarce and appropriate biosafety facilities are not easily accessible, but also where budget is a limitation and testing turnaround time is a major priority. The use of PKTP buffer could facilitate epidemiological surveillance in working environments, universities or schools, even in children where NPS testing is not ideal.

The calculated cost of the PKTP buffer is less than one US dollar per sample, which in conjunction with multiplexed reactions represents a clear advantage in favor of massive SARS-CoV-2 qPCR testing using saliva samples. The trade-off between diagnostic sensitivity (especially for samples with low viral load, which have been suggested to represent low-to-zero infection risk [25]), and the overall costs, implementation feasibility and testing capacity should be considered in different contexts aiming to optimize the use of scarce human and material resources.

## METHODS

### COVID-19 positive saliva samples

Saliva samples from COVID-19 patients were obtained at the National Institute of Respiratory Diseases (INER) in Mexico City, a tertiary referral hospital for COVID-19 during the pandemic. Participants included health-care personnel and patients with different disease severity. Samples positive for SARS-CoV-2 infection were selected according to viral load (Ct) values estimated with the CDC-validated protocol [20] (see details below). Saliva samples were collected by each participant by directly spitting into a sterile 15-mL tube. All samples were stored in 1.5 mL aliquots at -80 ºC in an appropriate biosafety level 3 facility. Fresh aliquots were used for all validation experiments to minimize freeze-thawing cycles. All donors provided written informed consent for the use of remnant samples for research. The protocol for patient enrolment and sample donation was revised and approved by the Institutional Review Board of the INER (protocol B12-20).

### PKTP buffer

PKTP buffer is composed of 1% Tween 80 (BP338-500 Fisher Scientific, Waltham, MA, USA), 5 mg/mL vinylsulfonic acid sodium salt solution (278416 Millipore Sigma, St Luis, MI, USA) and 200 µg/mL Proteinase K (Zymo Research, Irvin, CA, USA) in Ca- and Mg-free Dulbecco’s Phosphate Buffered Saline buffer (DPBS L0615-500 Biowest, Riverside, MO, USA). The saliva samples were mixed with PKTP on a 1:1, 1:2 or 1:3 PKTP to saliva ratio and heat-inactivated (10 min at 96 ºC) on a SimpliAmp Thermal cycler (Applied Biosystems, Waltham, MA, USA). 2 µL of the PKTP-inactivated sample were used for single or multiplex RT-PCR reactions. PKTP buffer was freshly prepared for all experiments except for the stability tests.

### RT-qPCR reaction

For singleplex and multiplex reactions, we used StarQ OneStep RT-qPCR kit (Genes2Life, Irapuato, Gto, Mexico) according to the manufacturer’s instructions. The final reaction volume was 10 µL, including 2 µL of StarQ Buffer 5X, 0.2 µL of StarQ Enzyme, 0.5 µL of probe mix containing 20X forward and reverse oligonucleotides and 2 µL of heat-inactivated saliva in PKTP buffer (2:1). Nuclease-free water was used as non-template control (NTC). The oligonucleotides and probes were used at concentration indicated in table 1 and the reactions were run in 384 well-PCR plates (Bio-Rad, Hercules, CA. USA). RT-qPCR were performed on a BIO-RAD CFX384 Real-Time System under the following conditions: 50 °C for 15 min, 95 °C for 2 min and 45 cycles of 95 °C for 15 s, 60 °C 30 s. For the CDC protocol, frozen saliva aliquots, with a previous positive COVID-19 test result were thawed on ice and 200 µL of sample were extracted with QIAMP Viral RNA Kit Cat. No. 52904 (Qiagen, Hilden, Germany) according to the manufacturer’s instructions. RNA was eluted in 50 µL of AVE buffer and 5 µL were used as template in a 20 µL reaction using the GoTaq Probe 1-Step RT-qPCR System Cat. No. A6120 (Promega, Madison, WI, USA). The N1-FAM, N2-FAM and hRP-FAM oligo/probe assays were from the 2019-nCoV CDC EUA Kit Cat. No.10006770 (IDT, Coralville, IA, USA). For comparison, PKTP reactions using the same samples of COVID-19 saliva were adjusted to 20 µL using 4µL of inactivated saliva-PKTP (2:1). Reactions from CDC protocol and PKTP inactivated samples were run together on Applied Biosystems™ 7500 Real-Time PCR System (ThermoFisher, Waltham, MA, USA). To mathematically adjust PKTP Ct values to compensate for dilution, 2.56 was subtracted from the PKTP values. Other oligo/probe assays where acquired from T4Oligo (Irapuato, Gto. Mexico) with HPLC purification.

**Table 1.**
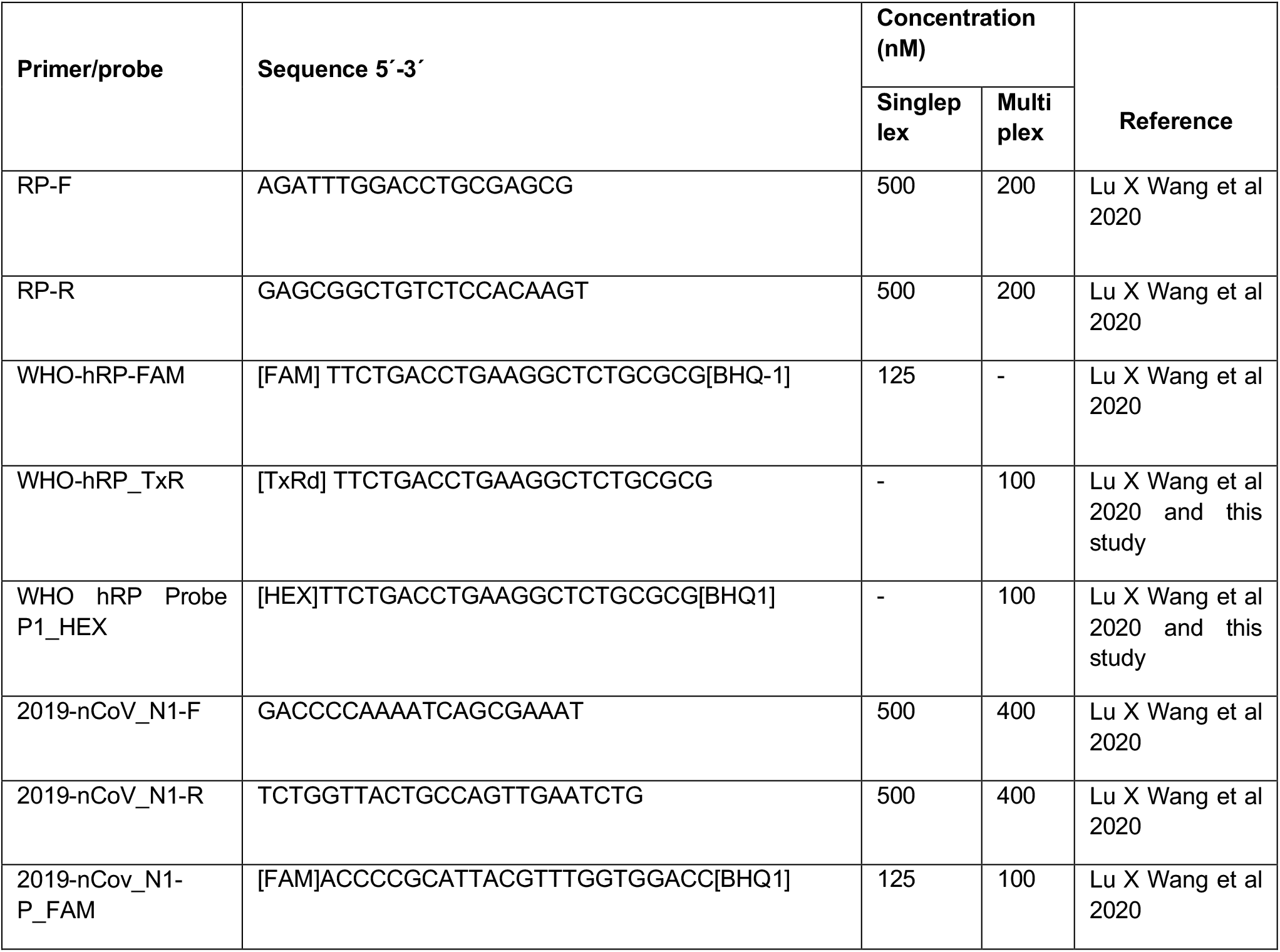
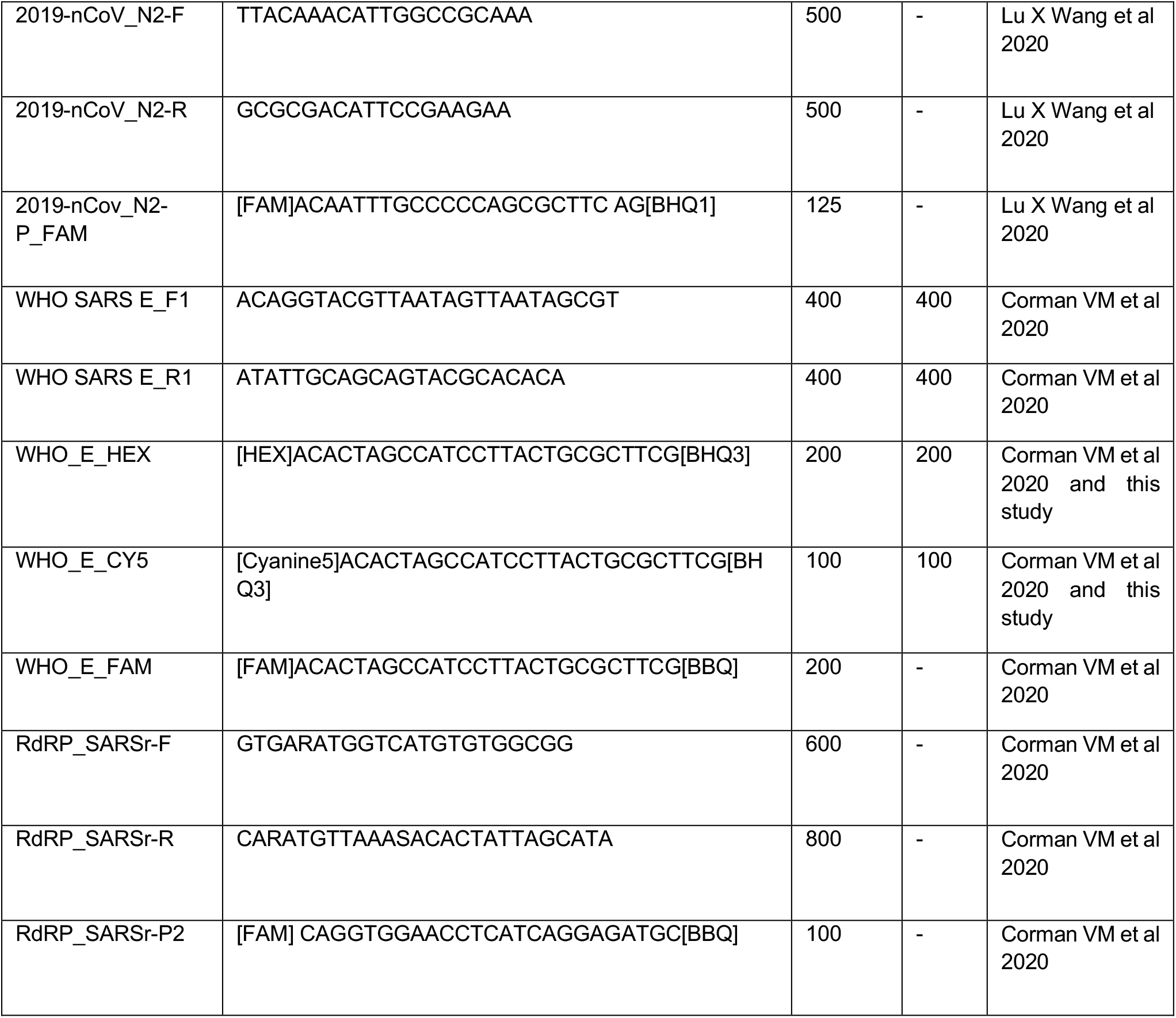
Oligonucleotides and probes used in this study.

### Self-collection of saliva samples

We provided a kit for self-collecting saliva containing a calibrated microcapillary tube cat # 2-000-050 (Drummond Scientific Company, Broomall, PA. USA), one 1.5 mL sterile microcentrifuge tube and one PCR tube with 10 µL of PKTP. Saliva was collected by directly spitting into the 1.5 mL microtube, and 20 µL of the sample were transferred into the PCR tube containing PKTP by participants using the microcapillary tube. Participants were instructed not to drink, eat, smoke or brush their teeth one hour before the test. The samples were heat-inactivated and stored for no more than 6 hours at -20 °C, except for stability tests.

### Total saliva RNA integrity test

COVID-19-negative saliva samples were divided into two, and each aliquot was incubated in either PBS or PKTP buffer on a 2:1 saliva:buffer ratio for 12 h at 4º C. After incubation, 200 µL of each sample were purified with QIAMP Viral RNA Kit Cat. No. 52904 (Qiagene, Hilden, Germany) according to the manufacturer’s instructions and subjected to electrophoresis in a 2100 Bioanalyzer using an RNA Pico kit Cat. No. 5067-1513 (Agilent, Santa Clara, CA, USA)

### Statistical Analysis

All variables were tested for normal distribution (Shapiro-Wilk test) and homogeneity of variance (Levene test). Parametric tests (ANOVA, Pearson’s Product Moment Correlation) were performed as indicated, since data met the necessary assumptions. All statistical analyses were undertaken in GraphPad Prism 9.0.1 (1992-2012 GraphPad Software, Inc) or the R statistical software version 3.6.3, using the PerformanceAnalytics package (V 2.0.4) for Pearson Correlation analysis {Brian G. Peterson, 2020 #28}

## Data Availability

All data produced in the present study are available upon reasonable request to the authors

## AUTHOR CONTRIBUTIONS

BBG, SGM, NCD and DMLS conducted the experiments. ASA, BBG, SGM, NCD, SAR and VJV performed data analysis and figures. BBG, SGM, NCD, ASA, GSMO, FRT, SAR and VJV conceptualized study design. FRT, SAR and VJV oversaw the study and funding. All authors contributed to writing the manuscript and approved its final version for submission.

## ACKNOWLEDGMENTS

This work was supported by the Secretaria de Educación, Ciencia, Tecnología e Innovación (CDMX) by grant SECTEI/277/219 to VJV. BBG (CVU: 1005898) was supported by Consejo Nacional de Ciencia y Tecnología (CONACyT) and SECTEI fellowships during his master studies. Sample collection and processing at INER was supported by a grant from CONACyT; Fondo FORDECyT-PRONACES to SAR. We acknowledge the technical support of Laura Ongay-Larios, Minerva Mora-Cabrera and Guadalupe Codiz-Huerta from the UBM, IFC, UNAM. Funding agencies had no role in study design, data analysis or interpretation of the results.

